# Challenges in using data on fathers/partners to study prenatal exposures and offspring health

**DOI:** 10.1101/2023.08.08.23293816

**Authors:** Kayleigh E Easey, Apostolos Gkatzionis, Louise AC Millard, Kate Tilling, Deborah A Lawlor, Gemma C Sharp

## Abstract

**Introduction:** Paternal exposures (and other non-maternal factors) around pregnancy could have important effects on offspring health. One challenge in research of paternal effects is that study samples with data on partners are usually a subgroup of those with data on mothers, which could introduce selection bias and limit generalisability of the findings. Here, we use maternal and father/partner data on prenatal behaviours to explore the extent to which selection in partner analyses might bias findings.

**Methods:** We characterise the availability of data on father/partner and mother health behaviours (smoking, alcohol consumption, caffeine consumption and physical activity) in the prenatal period from three UK cohort studies: the Avon Longitudinal Study of Parents and Children (ALSPAC), Born in Bradford (BiB) and the Millennium Cohort Study (MCS). We assess the extent of sample selection in these cohorts by comparing the characteristics of families where the father/partner does and does not participate. Using the association of parental smoking during pregnancy and child birthweight as an exemplar, we used simulations to explore the extent to which missing father/partner data may bias estimates.

**Results:** In all three cohorts, data on prenatal health behaviours of fathers/partners were less detailed and collected at fewer timepoints than data on prenatal health behaviours of mothers. Partners of mothers who had a lower socioeconomic position were less likely to participate. Estimates of the association between maternal smoking and offspring birthweight were similar in samples with and without participating partners in all three cohorts. In simulations based on ALSPAC data, there was little evidence of selection bias in associations of maternal smoking with birthweight, and although bias was observed for father/partner smoking, its magnitude was relatively small.

**Discussion:** Using real and simulated data, we show that bias due to selected recruitment of partners into ALSPAC, BiB and MCS had a relatively small impact on estimates of the effects of maternal and partner smoking on offspring birthweight. In other applications, the impact of selection bias will depend on both the analysis model and the selection mechanism. We have shown how to use a simulation study to assess that and recommend that applied researchers working with partner data use simulations and other sensitivity analyses to assess the robustness of their results.

## Introduction

The Developmental Origins of Health and Disease (DOHaD) hypothesis posits that environmental factors experienced in early life can have long-lasting influences on health and risk of disease. Much of DOHaD research is conducted on prospective, longitudinal (birth) cohort studies, with a strong focus on the effects of the intrauterine environment influenced by maternal exposures during pregnancy (1–4). These maternal pregnancy exposures are most commonly hypothesized to affect child health through direct influences on the developing fetus *in utero* (although transmission of epigenetic modifications via the maternal germline have also been studied (5)). Repeated measurements of maternal characteristics around pregnancy are often collected in cohort studies. This enables researchers to test the association of maternal exposures around pregnancy with offspring outcomes collected prospectively.

Increasing evidence suggests that father/partner exposures (and other non-maternal factors) around the time of pregnancy could also have important effects on offspring health (6–8). There are several potential mechanisms through which this can occur – either directly (e.g., for biological fathers, via germline transmission of epigenetic modifications) or indirectly via the mother (e.g., for any partner, through mothers passive smoking, or partners support that helps the mother quit smoking or drinking alcohol) (6, 9–11) (Figure 1). However, for research measuring paternal exposures around pregnancy, there is a lower breadth (fewer variables measured) and depth (simpler categories, collected less frequently) of available data. This can contribute to a lower quantity of published papers measuring paternal pregnancy effects (2, 8). Smoking in pregnancy for example, is a key maternal exposure known to cause lower birthweight in offspring (12, 13), yet studies seldom measure fathers/partners smoking behaviour in the same detail.

**Figure 1.**
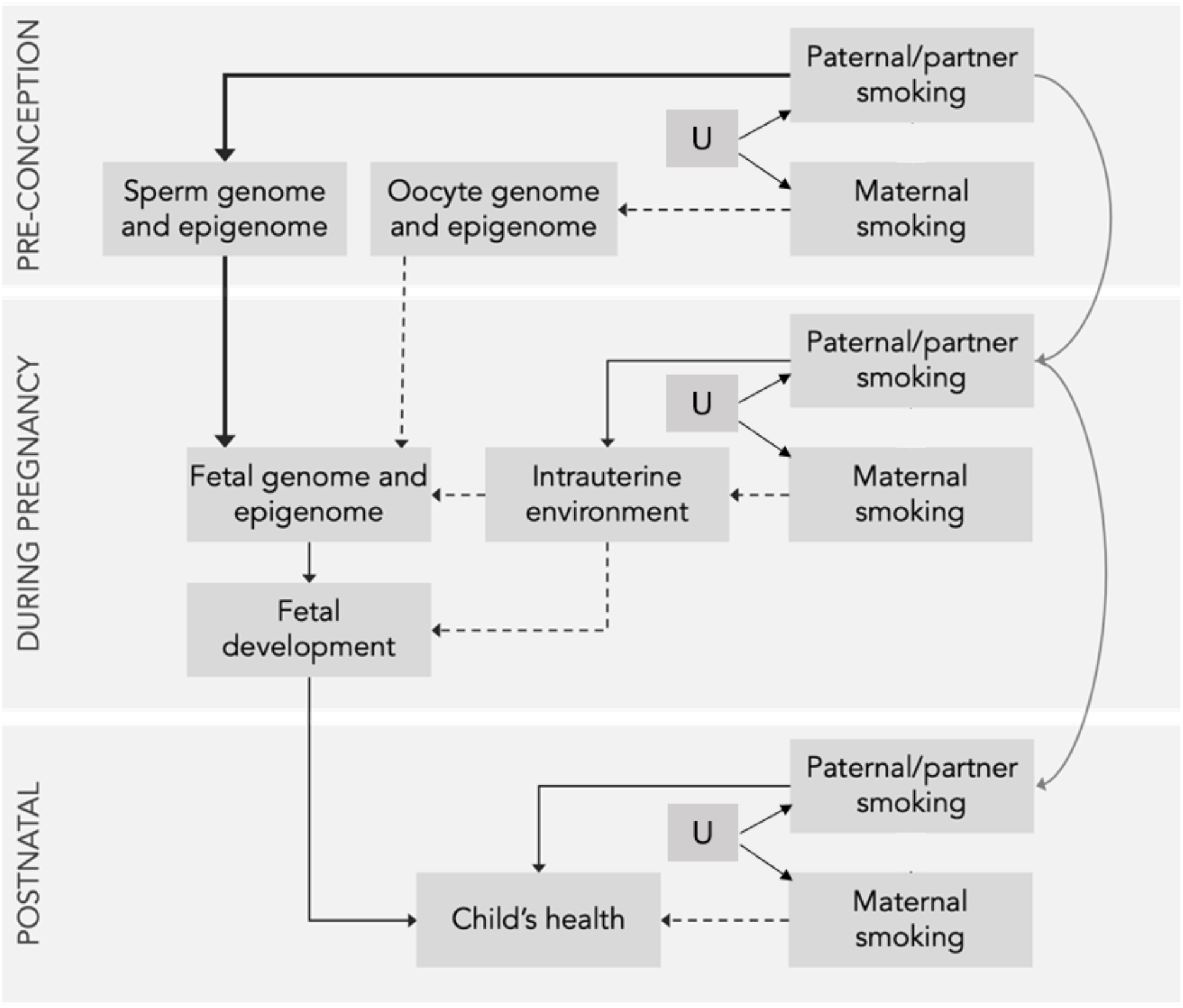
Potential pathways through which father/partner smoking could affect child health. Paternal/partner effects not via the maternal exposure are shown with solid arrows, and effects via maternal exposure are shown with dashed arrows. The thicker solid arrows highlight pathways that are only relevant to fathers/partners who are genetically related to the child (i.e., biological fathers). All other pathways are relevant to all partners. In these models there are unmeasured confounders, of which there will many and therefore represented by U. For simplicity, arrows between maternal smoking at different time periods are omitted, and likewise for the confounder.

The paucity of available partner data limits both the range of partner exposures in the prenatal period that can be studied and the ability to harmonise measures between parents and cohorts. The focus on maternal exposures over and above partner exposures in pregnancy could be argued to be due to a lack of availability of partner data in human cohort studies. However, a review of studies concerned with DOHaD, showed that lack of research interest in partner effects extends to animal studies too: only 10% of animal studies included partner measures, despite male animals being widely available for research (2). In some situations, there may be good reason for assuming maternal exposures are likely to have a stronger effect, but without exploring the impact partner exposures may have on offspring outcomes, we are ignoring an important pathway, as well as potentially reinforcing assumptions that it is only maternal behaviours during pregnancy that are important (1). In addition to investigating the impact partner prenatal behaviours have on child health, the collection of quality data on partners would improve analyses of maternal effects by being able to conduct negative control analyses using a partner exposure. Partner negative control studies are a method used to explore if maternal effects may be biased by residual confounding (14–16), and account for and block pathways of association from the partner exposure. Furthermore, in general, women are more likely than men to participate in research when invited, and in follow-up after pregnancy mothers are more likely to be the point of contact and respond to questionnaires about their children or bring them to research clinics that collect measurements and biological samples (17, 18). This results in an imbalance in the availability and completeness of data on mothers compared to partners.

We summarise available mother and partner data on four health behaviours (smoking, alcohol, caffeine, physical activity) around the time of pregnancy in three UK cohort studies – the Avon Longitudinal Study of Parents and Children (ALSPAC), Born in Bradford (BiB) and the Millennium Cohort Study (MCS). Using ALSPAC data, we conduct simulations to explore the extent to which missing partner data (hence selection into analytic subsamples) can bias estimates, using the association of parental smoking during pregnancy and child birthweight as an example. Using data from all three cohorts, we estimate the associations of maternal smoking with offspring birthweight in families with and without participating partners, to explore the “real life” impact of partner sample selection on estimates of maternal effects.

## Methods

For simplicity throughout the remainder of the paper we use the term ‘partner’ to include biological fathers and all other partners. In this paper, our aim was to describe differences in partner, compared with mother, recruitment into birth cohorts and to use simulation and real data to explore the potential bias this might introduce to estimating partner effects and in negative control analyses.

### Cohorts

ALSPAC, BiB, and MCS are UK-based prospective, ongoing multigenerational cohorts that recruited women and their partners during pregnancy (ALSPAC and BiB) or via household contact where there was a registered birth within a one-year set period (MCS). They include different populations (by geography and year of recruitment) and have different data collection and follow-up protocols, as described for each below. They also used different approaches to obtain partner data, and therefore understanding selection bias they may include would have a large impact for future research. These cohorts also have contrasting data collection and follow-up procedures, enabling us to investigate how the impact of selection differs across them.

### Avon Longitudinal Study of Parents and Children (ALSPAC)

ALSPAC recruited pregnant women residing in Avon, UK with expected dates of delivery between 1^st^ April 1991 and 31^st^ December 1992 (19, 20). The initial number of pregnancies enrolled was 14,541 pregnancies resulting in 14,062 live births, and 13,988 children who were alive at 1 year of age. Participants have been regularly followed up through clinic visits and questionnaires. Ethics approval for the study was obtained from the ALSPAC Ethics and Law Committee and the Local Research Ethics Committees. The study website (http://www.bristol.ac.uk/alspac/researchers/our-data/) contains details of all the data that is available through a fully searchable data dictionary and variable search tool. For the purposes of this study, we selected the first child from each cohort mother (n=14,472). Pregnant mothers were posted questionnaires at 18 weeks’ gestation to give to their partner to answer. If partners agreed, they were subsequently enrolled in the study. If mothers responded that they either had no partner, or that her partner did not want to take part then no further partner questionnaires were sent. Some information on partners was also collected by maternal report. For further details on the cohort profile, representativeness, and phases of recruitment, see articles by Boyd and colleagues (20); Fraser and colleagues (19) and Northstone and colleagues (21).

### Born in Bradford (BiB)

BiB recruited pregnant women residing in the town of Bradford, in the north of England (UK) between 2007 and 2011 (22). After being provided with initial study information at their first antenatal clinic appointment, pregnant women were largely recruited at a routine oral glucose tolerance test (OGTT) at 24-28 weeks’ gestation that all pregnant women in Bradford at the time of the study were invited to, unless they had pre-existing diabetes, which is very unusual and contrary to NICE guidelines (23). Around 80% of pregnant women in Bradford (without pre-existing diabetes) attended for an OGTT, and about 80% of these participated in BiB. This resulted in the recruitment of 12,453 women with 13,776 pregnancies and 13,740 live births (22). Ethical approval for BiB was obtained from the Bradford NHS Research Ethics Committee. We excluded mothers who had not completed the mother questionnaire at recruitment (i.e., those who completed it later in pregnancy or postnatally), giving a sample size of 11,395. For the purposes of this study, we randomly selected one child from each cohort mother. Mothers were not asked to report partners behaviours. Partner data was collected directly via self-report, through invitations to participate if they attended the OGTT or subsequent antenatal appointments with the pregnant study mother, or postnatally when a member of the study team visited the family home during follow-up visits.

### Millennium Cohort Study (MCS)

MCS recruited parents of children recorded as a live birth in the United Kingdom between 1^st^ September 2000 and 31^st^ August 2001 (for England and Wales), and between 24^th^ November 2000 and 11^th^ January 2002 (for Scotland and Northern Ireland) (24). To be eligible, the children had to be alive and resident in the UK at age 9 months, resulting in ∼19,000 children. Eligible children were identified using government child benefit records, which has almost universal coverage. Certain subgroups were intentionally over-sampled (designed for a disproportionate representation of families living in child poverty: children living in disadvantaged areas, children of ethnic minority backgrounds, and children growing up in the smaller nations of the UK), to be nationally representative of the UK population. In the first sweep, 18,827 children from 18,552 families were recruited. Ethical approval for the study was obtained from an NHS Research Ethics Committee (MREC) and informed consent was obtained from parents for their children’s participation. For the purposes of this study, we excluded 55 children where the mother was not the biological mother, and randomly selected one child from each pregnancy (*n*=18,241).

Within a household both mothers and partners were invited to take part via interview, at which information on non-resident parents were also collected, via the parent who was resident. The first sweep interviewed both mothers and (where resident) fathers/other parents of infants included in the sample. Non-resident parents were invited to take part within the later sweeps.

### Extraction and harmonisation of data

#### Paternity and maternity status

In ALSPAC and BiB, all cohort mothers were recruited during pregnancy and were biologically related to their children. In ALSPAC, mothers were asked whether the partner they had identified to be invited to the study was the biological father. In BiB, cohort mothers or their partner were not asked whether the partner was the biological father. In MCS, as described above we excluded families where the household main respondent reported that they were not the biological mother. Co-resident partners in the household were asked to report their relationship to the cohort child, including whether they were the child’s biological parent.

#### Definition of partner participation

For each cohort, we created a binary variable to describe the participation of the partners of cohort mothers. Partners were defined as “participating” if they had provided any data via self-report questionnaire or interview in the initial stages of the studies. In ALSPAC, participating partners had to have completed and returned at least one item from the first three questionnaires given to partners around pregnancy, after enrolment through the pregnant mother. In BiB, participating partners had to have returned the baseline questionnaire, sent to partners during or shortly after the study pregnancy. In MCS, participating partners had to have been interviewed during the first sweep interview.

#### Partner and maternal health behaviours

For each cohort, we described availability of measures of parental smoking, alcohol consumption, caffeine consumption and physical activity in the prenatal period. Exposure timepoints of interest were the three months prior to pregnancy (preconception), any time during pregnancy, and in each of the three trimesters separately. For smoking, we generated binary (any/none) variables to describe smoking behaviour of both the mother and partner for each timepoint. More details are provided in the Supplementary Methods.

#### Other partner, maternal and birth characteristics

Across the three cohorts, we derived and harmonised (where possible) variables describing parent and birth characteristics. Self-reported measures were maternal age in years at conception (BiB) or delivery (ALSPAC and MCS); partner age during pregnancy (BiB) or delivery (ALSPAC and MCS); highest level of education; mother and partner ethnicity (white European or Black/Asian/mixed/other); maternal pre-pregnancy body mass index (BMI). Mother reported co-habitation status (partner lives with mother during pregnancy vs not), marital status (married vs not married). Mother-report or birth record of baby’s birthweight (grams); gestational age at delivery (weeks). To compare differences in participation across different characteristics on the same scale, we categorised continuous and ordinal measures: parental age (young <=21 years; high >=35 years); maternal pre-pregnancy obesity (low BMI <30kg/m^2^; high BMI >= 30) baby’s birthweight (low <2.5kg; high >4.5kg); preterm birth (<37 weeks vs >=37 weeks); parental education (university degree or higher vs any lower qualifications/no qualifications).

Because partner data is not generally available for partners that do not participate, we included mother and birth characteristics (maternal age, obesity, ethnicity, education, co-habitation status, marital status, birthweight, gestational age at birth).

#### Hypothesised selection bias due to partner participation

We hypothesised that differences in socioeconomic position (SEP) and parental health behaviours between families with participating partners and those without could introduce selection bias. The directed acyclic graph (DAG) in Figure 2 illustrates how missing partner data might introduce selection bias in studies of the effects of partner or maternal smoking (exposure) on child birthweight (outcome). This DAG shows our assumptions, including that partner SEP, smoking and general health influence partner participation, which could result in spurious associations between these in analyses that do not account for this selection into the study.

**Figure 2.**
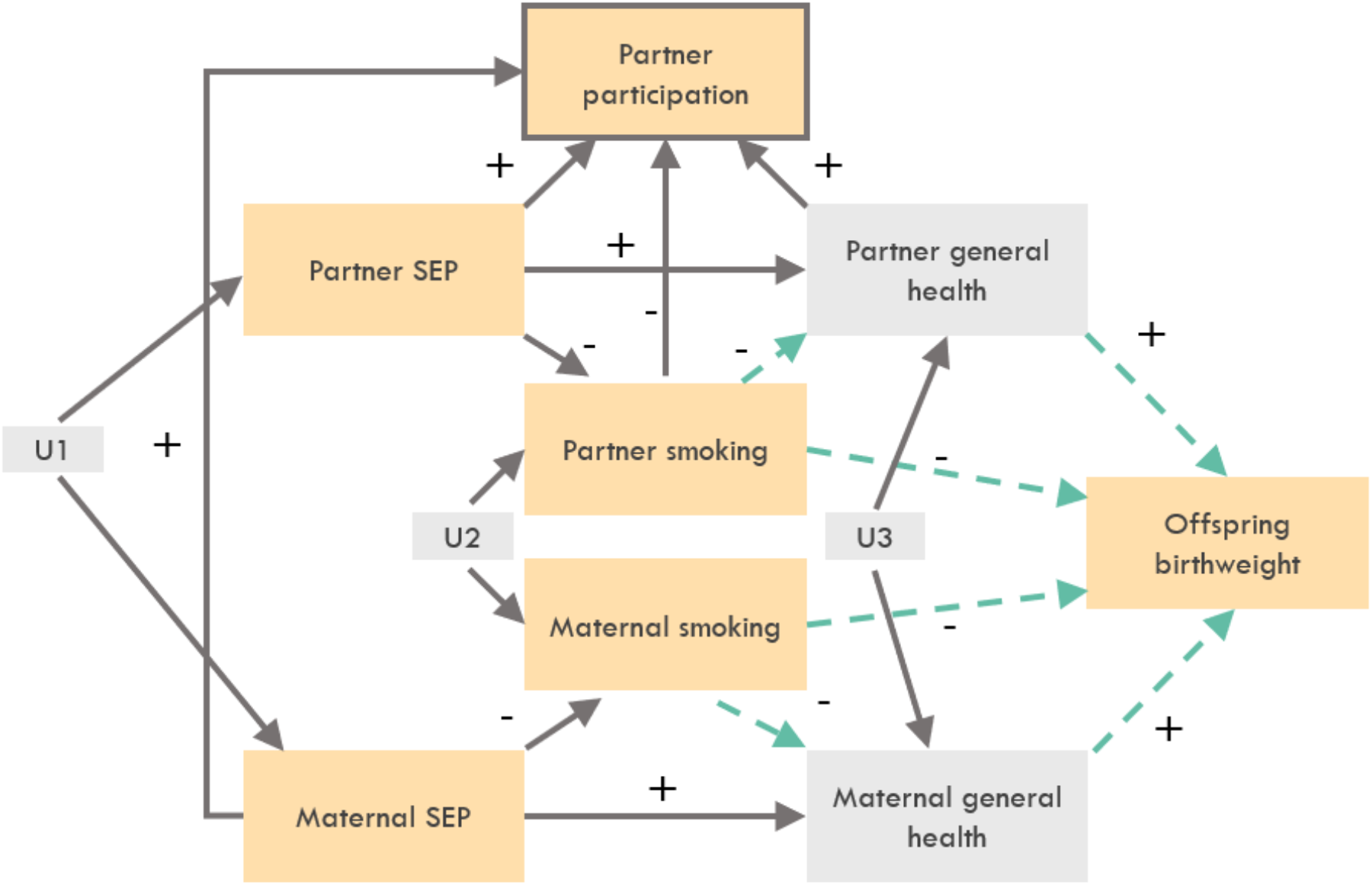
Directed acyclic graph (DAG) illustrating how missing partner data (incomplete partner participation) might introduce selection bias. Grey shading indicates variables that are not usually measured in real life studies (but were simulated). Dashed green arrows indicate the main effects being estimated and other arrows indicate causal effects. A box is drawn around partner participation to indicate that it is conditioned on through selection.

### Descriptive analysis of cohort characteristics and sample selection

To characterise the availability of data on partner and mother health behaviours in the prenatal period, we summarised the availability (yes/no) of our health behaviours of interest and calculated the percentage of partners participating in each cohort.

To assess the potential for sample selection in these cohorts we used logistic regression to estimate the univariable association of family characteristics in pregnancy with partner participation. Where partner data had been provided by the cohort mother, we also compared partner participation across mother-reported partner’s age, ethnicity, education, and paternity status.

### Simulation study to explore the magnitude and direction of bias due to sample attrition

We conducted a simulation study to assess the impact of selection bias, using the association of maternal and partner smoking during pregnancy with child birthweight as an example. Our simulations were designed using the ADEMP framework (25) and were informed by the ALSPAC dataset, since there was more information on partner variables in ALSPAC than in the other two cohorts (Figure 3). The ADEMP formulation of our simulation study was as follows:

**Figure 3.**
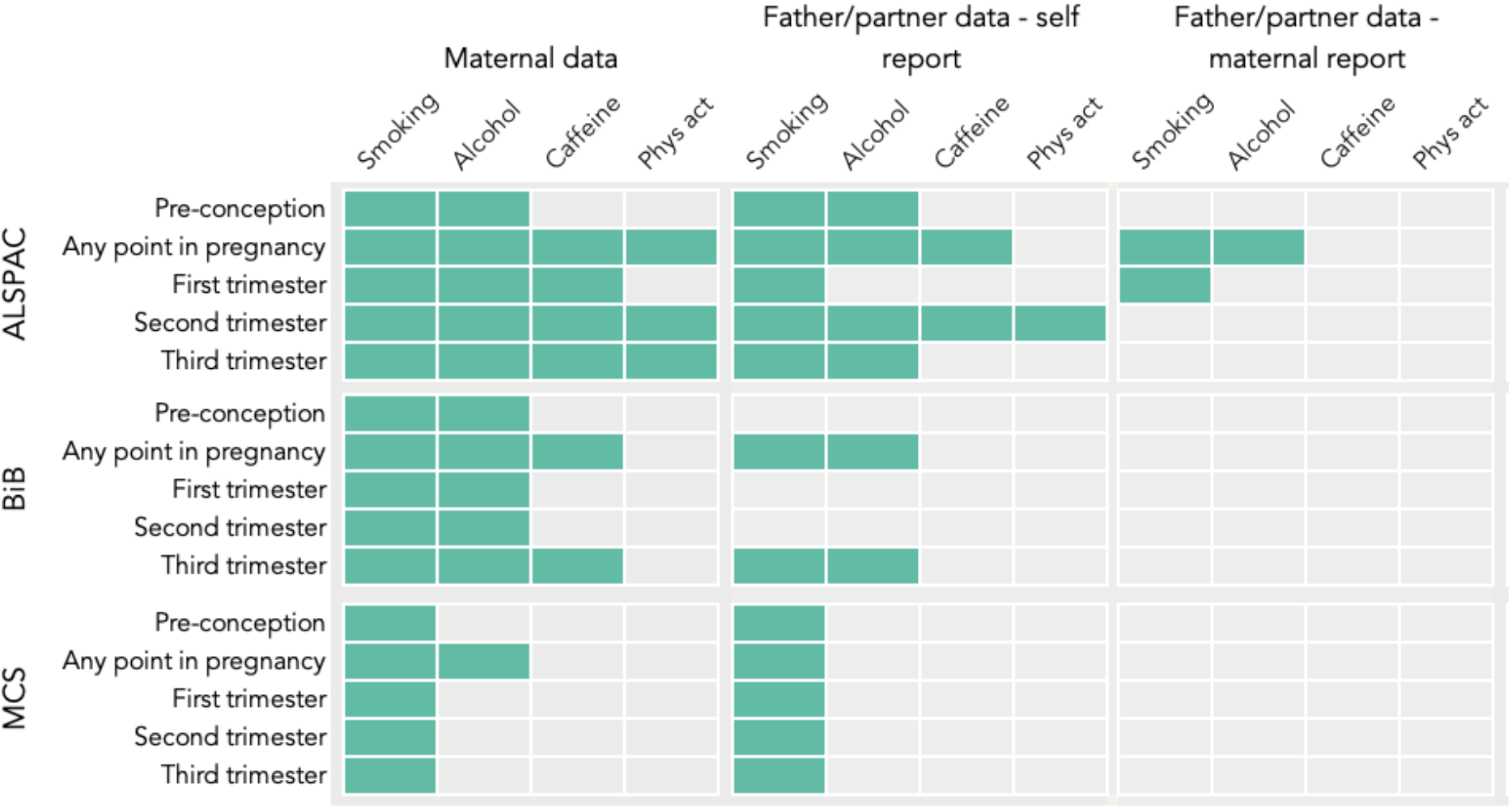
Availability of data on parental prenatal health behaviours in ALSPAC, BiB and MCS.

#### Aim

To assess the impact of selection bias on an ALSPAC-based analysis of the association between parents’ smoking and offspring birthweight.

#### Data-generating model

For our main simulation, we generated data on 15,000 individuals, according to the DAG in Figure 2. Associations between the variables in Figure 2 were estimated using ALSPAC data where possible and used to parameterize the simulations. Parental “general health” variables could not be measured directly in ALSPAC. Therefore, to estimate associations involving these variables we used body mass index (BMI) as an indicator of general health. Moreover, missing partner data on non-participating partners meant that the associations of partner SEP, smoking and health with partner participation could not be estimated directly. To specify the effects of these variables on participation in our simulations, we generated eleven simulation scenarios, which are summarised in Supplementary Table 1. In Scenario 1, we set the effects of partner health and smoking on partner participation equal to the observed associations of maternal BMI and smoking with partner participation in ALSPAC (0 and −0.81 respectively) and estimated the association of partner SEP with participation using data reported by the mothers about their partners. In Scenarios 2-6, we assumed no direct effect of partner smoking on participation and set the participation effects for SEP and BMI to be such that the odds ratio for participation per one category increase in SEP (coded as a categorical variable with four categories) and the odds ratio for participation per one standard deviation increase in BMI were equal to 1.5, 2, 5, 10 and 20 respectively. Some of these values are large, e.g., in Scenario 1, the odds ratio for participation per unit increase in partner SEP as reported by mothers was 1.58. Nevertheless, we chose to use these values to explore the impact of selection bias for a range of different strengths of association between partner traits and participation. In scenarios 7-11, we also assumed an effect of partner smoking on participation, with an odds ratio of 0.67, 0.5, 0.2, 0.1 and 0.05 respectively (see Supplementary Methods). Again, these values were selected to represent different magnitudes of association between partner smoking and participation (we used odds ratios lower than 1 since smoking is negatively correlated with participation in ALSPAC).

In addition to our main simulation described above, we conducted two more sets of simulations. In the first set of simulations, we modified the DAG of Figure 2 by including an unobserved variable acting as a common cause (confounder) of partner participation and birthweight. In the second set of simulations, we included an interaction between partner smoking and BMI in their effects on participation. Both of these modifications are likely to increase the magnitude of selection bias. In the first case, collider bias may be induced between partner smoking and the common cause of participation and birthweight. In the second case, it has been suggested in the literature that selection bias can have a bigger impact in analyses where the exposure and outcome interact in their effects on study participation (21, 22); here, an interaction between smoking and birthweight would not be expected, since partner participation in ALSPAC was determined before childbirth therefore birthweight could not be a cause of participation, so instead we simulated an interaction between partner smoking and partner general health. By conducting these additional simulations, we sought to assess the sensitivity of the results of our main simulation to these two aspects of the data generating model (outcome-participation confounding and interactions in the model for participation).

#### Estimands

The associations of maternal and partner smoking with offspring birthweight, adjusted for each other and for both parents’ SEP. We did not adjust for parents’ general health, as this variable is not directly observed in ALSPAC.

#### Methods

Estimates were obtained from a linear regression model, fitted only using data on families with participating partners, with offspring birthweight as the outcome and maternal smoking, partner smoking, maternal SEP and partner SEP as covariates.

#### Performance measures

Absolute and relative bias, model-based standard errors, empirical coverage of 95% confidence intervals, and bias-eliminated coverage. Results were averaged across 1000 repetitions of the simulation.

We refer to the Supplementary Methods for more details about our simulation design and its ADEMP formulation.

### Applied analysis of potential selection bias

To assess the extent of selection bias in real data, we conducted linear regression analyses and compared the effect estimates of maternal smoking on birthweight in ALSPAC, BiB and MCS using the total sample and samples stratified by partner participation status, both with and without adjustment for partner smoking.

Analyses were performed in R version 4.0.3 or Stata version 15, and code is available at [https://github.com/agkatzionis/Parental-smoking-and-birthweight.git].

## Results

### Partner participation

Of total pregnancies in each cohort there were the following with a cohort participating partner: In ALSPAC, of 14,472 pregnancies, 12,997 (78%); In MCS, of 18,241 pregnancies, 13,145 (71%); In BiB, of 11,538, 3,131 (27%).

### Available data on parental prenatal health behaviours

In all three cohorts, data on partner prenatal health behaviours were less detailed and collected less frequently than data on maternal prenatal health behaviours (Figure 3).

### Family characteristics by partner participation

Figure 4 shows the pattern of association between family characteristics and partner participation. Consistently across all cohorts where data was available, mothers were more likely to have a participating partner if the woman was white, had a degree, or was cohabiting with her partner before the birth of the child. Families where babies were born preterm or with a low birthweight were less likely to have a participating father/partner.

**Figure 4.**
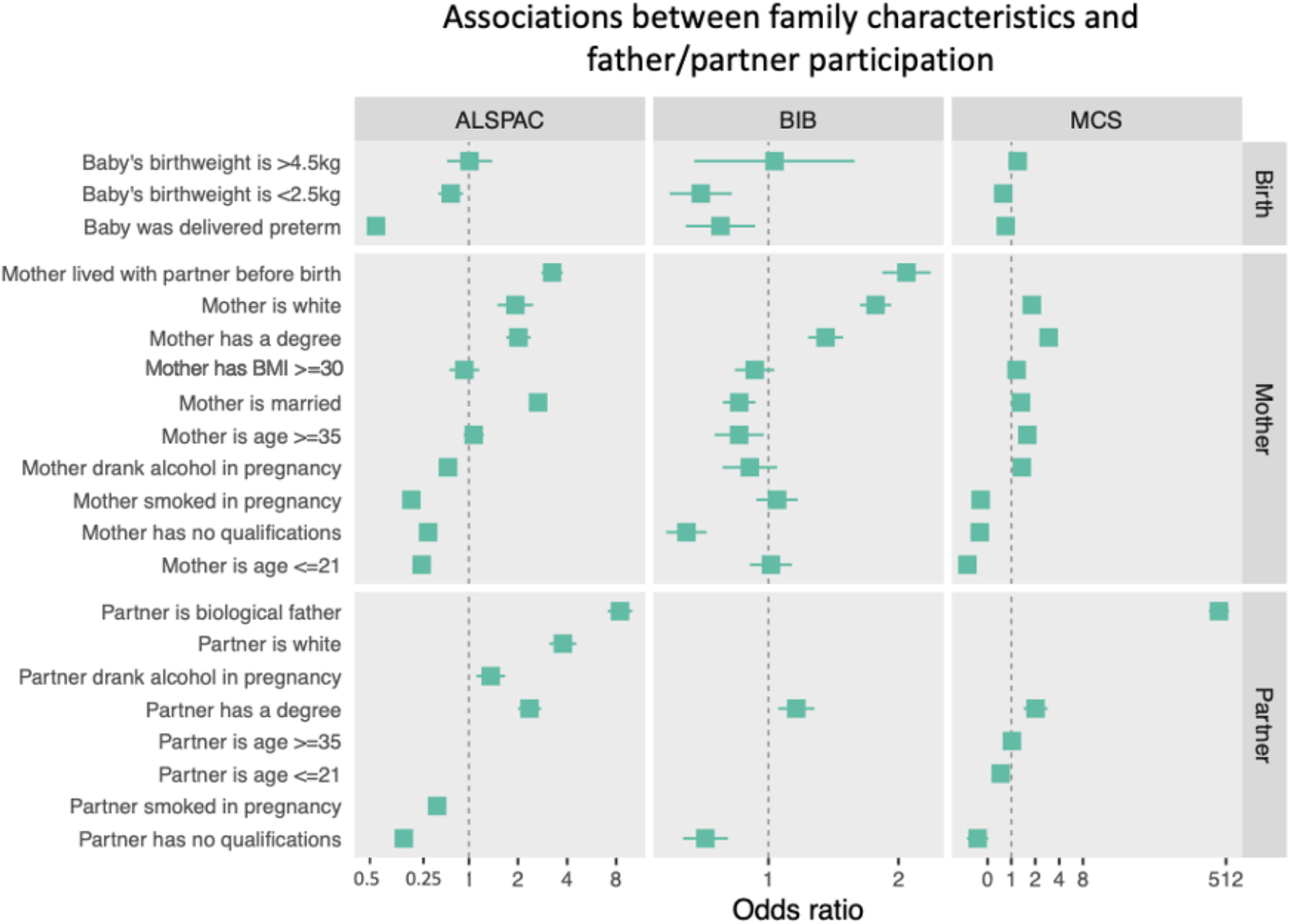
Association between family characteristics and participation of the partner in the cohort study. Squares represent the odds ratio of participation for each binary characteristic; horizontal lines show the 95% confidence intervals.

Maternal report of partner measures showed that partners were more likely to participate if they were the biological parent, were white, had consumed alcohol during pregnancy, and had a degree. Those who were smokers or had no qualifications were less likely to participate. Other family characteristics, such as maternal alcohol consumption during pregnancy and parental age, had less consistent associations with partner participation across the three cohorts (Figure 4).

In all cohorts, higher SEP and being white were associated with higher rates of partner participation. Whereas higher SEP was associated with lower rates of smoking, being white was associated with higher rates of smoking (Figure 4), (97% white in ALSPAC; 75% white in MCS; 40% white in BIB). A large proportion of participants in BiB (51%) identify their ethnicity as South Asian. Rates of smoking in this group are lower than in the white group, particularly rates of maternal smoking during pregnancy (3% in South Asian mothers, 34% in white European mothers; 30% in South Asian partners, 34% in white European partners).

### Simulation study to explore the magnitude and direction of bias due to sample attrition

Figure 5 and Supplementary Table 2 show the results of the simulation study to assess the extent to which missing partner data could introduce selection bias. For the associations of maternal smoking with offspring birthweight there was little evidence of selection bias in any of the eleven scenarios, with estimates being close to the true value of the maternal effect on smoking in our simulations (relative bias less than 1%). For partner associations, there was some evidence of selection bias in Scenarios 9-11, where all three partner traits (SEP, smoking and health) had strong effects on participation. The presence of bias can be justified theoretically, according to the theory of causal diagrams: using only data on families with participating partners induces bias through the path “Partner smoking → Partner participation ← Partner Health → Birthweight”. In our simulations, the bias acted in a positive direction. The direction of bias can also be justified theoretically by noting that partner smoking leads to reduced participation while better partner health increases participation; therefore, among participating families, the association between partner smoking and general health will be overestimated (26) and so will the association of partner smoking with offspring birthweight. However, our results show that the magnitude of bias was small: across our 11 scenarios, the worst-case absolute bias was 0.02 and the worst-case relative bias was 12.6%. Similarly, the empirical coverage of 95% confidence intervals attained nominal levels in most simulation scenarios; in the worst-case scenario (partner smoking, Scenario 11) the empirical coverage was 83.6%. The bias would be eliminated if it was possible to adjust the analysis for partner general health, as we demonstrate in Supplementary Table 3.

**Figure 5:**
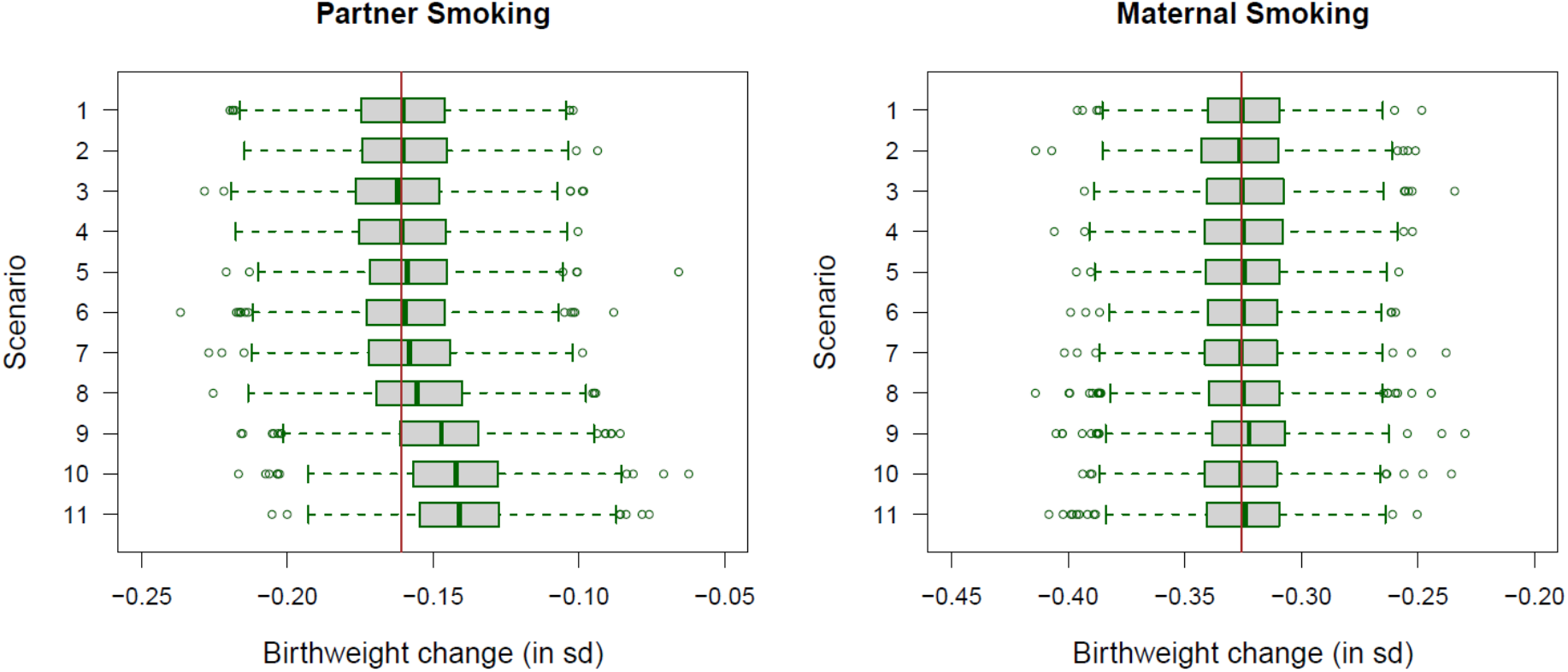
Boxplots summarising the results of our simulation study. Estimated associations of partner (left) and maternal (right) smoking with standardised birthweight (standard deviation increase in birthweight for smokers compared to non-smokers). Estimates are obtained from a joint analysis of both maternal and partner smoking, and are also adjusted for both parents’ SEP. A brown line indicates the true value of the parameters to be estimated. The results are provided in numerical form in Supplementary Table 2.

There was little evidence of selection bias for either maternal or partner associations in the scenario where partner effects on selection were set according to the observed associations with maternal traits in ALSPAC (absolute bias 0.001, relative bias 0.5% for paternal/partner estimates).

The results of the two additional sets of simulations are reported in Supplementary Tables 4 and 5. Adding a common cause of participation and birthweight increased the impact of selection bias on partner association estimates, although again there was little evidence of bias in maternal estimates. Even for partner estimates, the magnitude of bias remained small (relative bias less than 20% in all scenarios) and conclusions from the analysis were not seriously affected (both maternal and partner smoking were on average associated with reduced offspring birthweight in all scenarios). Similar results were obtained from the simulation with a smoking-general health interaction (worst-case relative bias 15.2% and empirical coverage 78.7%).

### Influence of missing partner data on partner-adjusted estimates of maternal effects using real (non-simulated) data

Figure 6 shows estimates of association between maternal smoking and offspring birthweight in ALSPAC, BiB and MCS (adjusted for partner smoking, and with and without adjustment for maternal ethnicity). We saw relatively stable effect estimates within cohorts regardless of whether we used the sample with or without participating partners, or adjusted or not for partner smoking. The exception was in BiB, where the association attenuated towards the null after adjustment for partner smoking.

**Figure 6:**
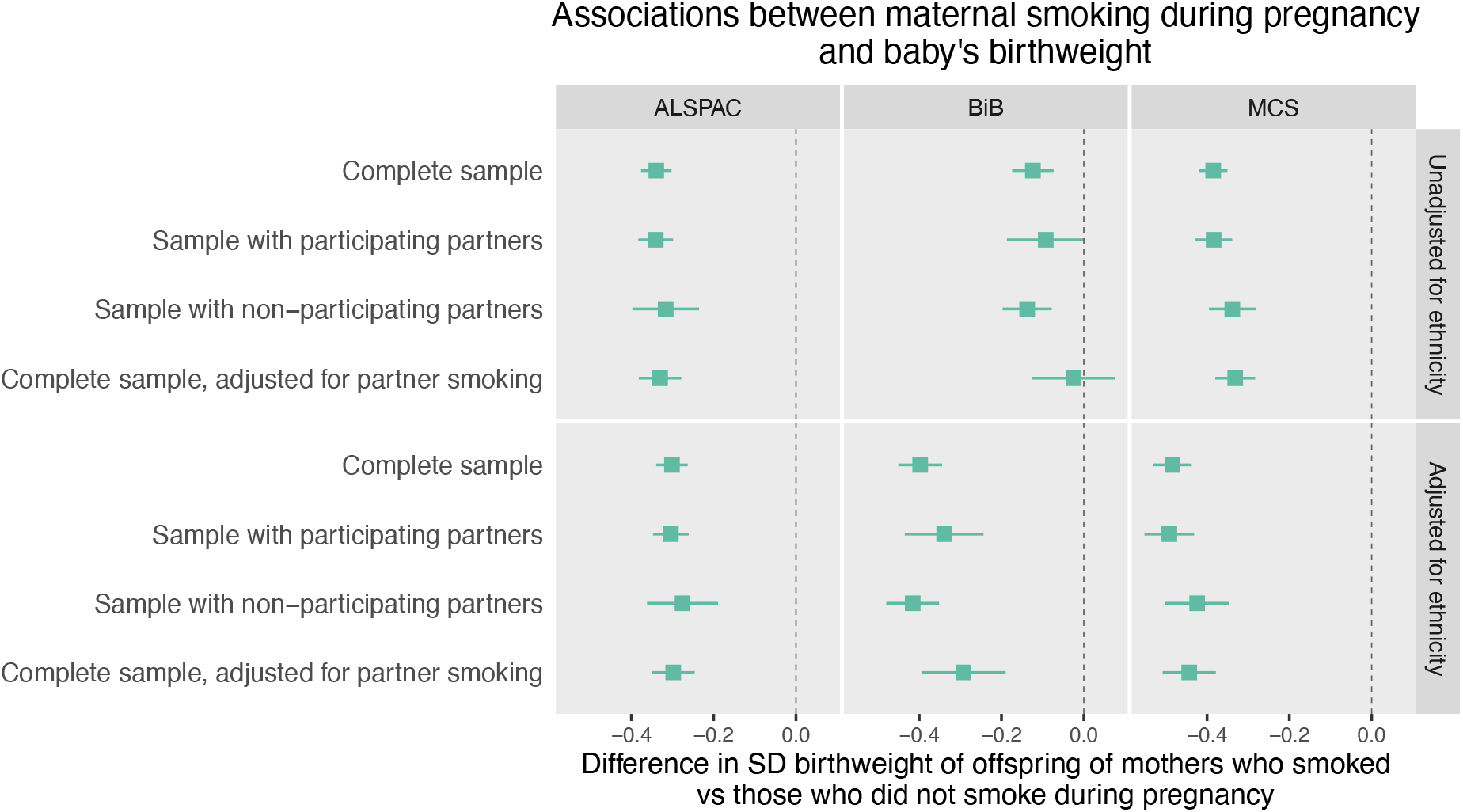
Effect estimates of maternal smoking during pregnancy on offspring birthweight (z-scores) in ALSPAC, BiB and MCS. Linear regression was run on either the complete sample, samples stratified by partner participation, or the sample with participating partners adjusted for self-reported partner smoking sample (the latter model restricts the sample to participating partners). Models were run unadjusted (top row) and adjusted (bottom row) for maternal ethnicity.

## Discussion

We have described the availability of data on partner prenatal health behaviours in three UK cohorts and explored how missing partner data might influence findings from studies of prenatal parental effects on child health. All three cohorts collected some self-report data from partners regarding health behaviours in the prenatal period. All four health behaviours (smoking, alcohol, caffeine consumption, physical activity) were collected at more time points for mothers compared to partners. All cohorts included at least some self-report data on smoking during pregnancy for both mothers and partners. Physical activity had the least amount of data and was only available in one cohort (ALSPAC), at three timepoints for mothers and only one timepoint for partners.

Partners who participate in cohort studies have been shown to have higher SEP compared to partners who do not participate (see (27) for a review). This underrepresentation of certain demographics may contribute to selection bias. We tested whether participating partners (only available through the mother’s participation) were systematically different from non-participating partners, using self and maternal report of partner characteristics (where we did not have self-reported data on non-participating partners), and also indirectly using maternal and birth characteristics, which are likely to be correlated with partner characteristics due to assortative mating, shared environments, and shared father-offspring genetics. Across all three cohorts we studied, pregnancies with participating partners were more likely to have a degree, be married and over 21 years old. This suggests that partners who did not participate in these cohorts were likely to have a lower SEP than those who did. These findings are in agreement with a previous study (28) that showed that working class fathers, less educated fathers and fathers from minority ethnic groups were underrepresented in the National Institute of Child Health and Human Development (NICHD) Study of Early Child Care. Moreover, babies of participating fathers were healthier with higher birthweights than babies of fathers who did not participate in that study (28).

In our real-data analyses, estimates of the effects of maternal smoking on offspring birthweight did not differ substantially when using the complete (unselected) sample, the sample where partners participated, or the sample where they did not. This was the case in all three cohorts and suggests that, although adjusting for missing partner data can introduce selection bias, in the examples we investigated, the biasing effect appears to be small. This was confirmed by a simulation study informed by the ALSPAC dataset. The importance of mutually adjusting maternal and partner exposures in these negative control analyses has been demonstrated (29, 30) and it is possible that lower recruitment of partners may then result in bias of both maternal (real) and partner (negative control) results; in our simulations, however, only the latter was observed.

There were relatively stable effect estimates within cohorts for if we used the sample with or without paternal participation or adjusted for paternal smoking, except for within BiB where the association attenuated towards the null after adjustment for paternal smoking, this is likely to be because of confounding by ethnicity. In all cohorts, being white and a higher SEP were associated with higher partner participation, however, higher SEP was associated with lower rates of smoking, and being white was associated with higher rates of smoking. Because BiB is ethnically diverse, the association between maternal smoking and birthweight may be confounded by ethnicity and the results are therefore presented both unadjusted and adjusted by ethnicity. This confounding relationship has little effect on the estimates generated in ALSPAC and MCS because participants in these cohorts are mostly white (97% white in ALSPAC, 75% white in MCS). However, only 40% of BIB are white and a large proportion of participants (51%) identify their ethnicity as South Asian. Rates of smoking in this group are lower than in the white group, particularly rates of maternal smoking during pregnancy (3% in South Asian mothers, 34% in white European mothers; 30% in South Asian partners, 34% in white European partners). Therefore, ethnicity (or rather, cultural factors proxied by ethnicity) is a major source of variation in maternal smoking data in BiB.

The strengths of our study include the use of three large UK cohorts that are commonly used to study maternal, and increasingly, partner effects on offspring health (15, 31). The use of simulations alongside the real-data analysis allowed us to explore the effects of selection bias. The simulation parameters were, where possible, informed by ALSPAC data to make them more realistic. Limitations include not examining cohorts from different countries which may have used different recruitment strategies, as well as having different cultures of research participation. Within the simulation study we used BMI as a proxy for general health, but the relationship between BMI and other variables in our analysis may not be the same as these variables’ relationship with general health. For example, while we would expect SEP and general health to be positively correlated, the SEP-BMI association that we observed in ALSPAC was negative (because individuals with a high SEP are less likely to have very high BMI). The aim of this paper however was to compare maternal-offspring associations with the equivalent partner-offspring associations. We also only explored a prespecified research question and a small set of scenarios (utilising the health behaviour, smoking, as most readily studied across cohorts), future research may seek to investigate using additional measures. Lastly, within non-participating partners, we were unable to distinguish between partners who declined to participate and partners of mothers who stated that they did not have a partner. There may be differences in characteristics between these partner groups which could impact any bias.

We agree with others (6, 28) that more studies of partner effects (in their own right) and studies of maternal effects that adjust for partner data, are needed. However, as with all epidemiological studies, care should be taken when designing and interpreting results from studies that use partner data. Based on our findings here, our recommendations for future data collection within new cohorts and for studies including of the effects of prenatal parental health behaviours are as follows. Firstly, efforts should be made to maximise partner recruitment where possible. Secondly, newly designed cohort studies should collect data with the same frequency and using the same questions in both partners and mothers. Third, maternal report of partner measures should also be collected for use where partner self-report data is unavailable, particularly as maternal report has been shown to be an accurate measure of partner behaviour in some circumstances (32). Fourth, data should be collected concerning paternity, because biological and non-biological partners affect offspring health through different mechanisms. Lastly, we would recommend that studies investigating the impact of prenatal exposures should, where possible, adjust maternal effects for partner data (and vice versa). We support the use of mutual adjustment to prevent bias from assortative mating (30).

Our study has highlighted discrepancies in the breadth and depth of cohort study data on the health behaviours of partners compared to mothers. Theoretically, missing father/partner data could introduce selection bias in studies of parental effects on child health. However, using real and simulated data, we have shown that any biasing effect of partner selection to birth cohort studies is likely to be small, although this may differ across cohort studies in different populations and with different selection mechanisms. We hope that our findings and recommendations encourage more research that collects and analyses data on partners.

## Data Availability

The study website (http://www.bristol.ac.uk/alspac/researchers/our-data/) contains details of all the data that is available through a fully searchable data dictionary and variable search tool. Access to the raw data is available at a cost by emailing

https://www.bristol.ac.uk/alspac/researchers/access/

## Acknowledgements and funding

ALSPAC: We are extremely grateful to all the families who took part in this study, the midwives for their help in recruiting them, and the whole ALSPAC team, which includes interviewers, computer and laboratory technicians, clerical workers, research scientists, volunteers, managers, receptionists, and nurses. This work was supported by the UK Medical Research Council and the Wellcome (Grant ref: 217065/Z/19/Z) and the University of Bristol provide which provide core support for ALSPAC. This publication is the work of the authors and GCS will serve as guarantor for the contents of this paper. This work was performed in the UK Medical Research Council Integrative Epidemiology Unit, jointly funded by the University of Bristol and the UK MRC. KEE and GCS were financially supported by an MRC New Investigator Research Grant awarded to GCS (grant code MR/S009310/1). GCS is also financially supported by the European Joint Programming Initiative “A Healthy Diet for a Healthy Life” (JPI HDHL, NutriPROGRAM project, UK MRC MR/S036520/1). AG, KT (MC UU 00032/02) and DAL (MC UU 00032/05) are supported by the Medical Research Council Integrative Epidemiology Unit at the University of Bristol. We are grateful to the Centre for Longitudinal Studies (CLS), UCL Social Research Institute, for the use of these data and to the UK Data Service for making them available. However, neither CLS nor the UK Data Service bear any responsibility for the analysis or interpretation of these data.

