## Supplementary Materials for "Challenges in using data on fathers/partners to study prenatal exposures and offspring health"

**Supplementary Methods**

*ADEMP framework for simulation study*

We used the ADEMP framework of Morris et al. (2019) to design our simulation study. In the next sections, we provide an outline of our simulation design according to the ADEMP framework.

*Aim*

The aim of our simulation was to assess whether selection bias will occur in an ALSPAC-based study of the effects of maternal and paternal smoking on offspring birthweight. Selection bias is suspected because paternal participation was optional in ALSPAC and there may be differences in the characteristics of non-participating fathers compared to participating ones.

*Data-generating model*

The design of our simulation study was based on the causal diagram on Figure 2 in the main part of the paper and was informed by the ALSPAC dataset. For our simulations, we used a sample of $N = 15000$ individuals, which is approximately the sample size of the ALSPAC dataset.

Socioeconomic position (SEP) was reported as a categorical variable in ALSPAC with four categories (0/1/2/3) and frequencies 20.2%, 44.5%, 22.4% and 12.9% respectively for each category. Higher categories represent individuals with better SEP. In our simulations, we generated two continuous variables, $L_M, L_P \sim N(0, 1)$, representing latent maternal and paternal SEP, and then used suitable cut-off points to convert these continuous measurements to categorical variables $SEP_M, SEP_P$ taking values 0/1/2/3 with the same frequencies as in ALSPAC. The continuous variables $L_M, L_P$ were correlated to ensure that $Cor(SEP_M, SEP_P) = 0.575$, which was the observed correlation between maternal and paternal SEP in ALSPAC.

Maternal and paternal smoking were generated as binary variables with smoking probabilities:

$$logit\ P(Smoking_M = 1) = -0.692\ SEP_M + 2.65\ U_1 + \log(0.46)$$

$$logit\ P(Smoking_P = 1) = -0.608\ SEP_P + 2.65\ U_1 + \log(1.92)$$

where $U_1$ is a confounding variable that models the correlation between paternal and maternal smoking patterns. The coefficients $-0.692$ and $-0.608$ were the observed effects of maternal and paternal SEP on smoking status obtained from logistic regression analyses in ALSPAC, while the confounding coefficient 2.65 was chosen so that the correlation between paternal and maternal smoking status was the same as in ALSPAC. Finally, the intercept terms were chosen so that the smoking frequencies were approximately 30.6% for mothers and 48.5% for fathers, which are again the values observed in the ALSPAC dataset.

As discussed in the main part of our manuscript, we used BMI as a proxy for general health when generating the simulations. BMI measurements in ALSPAC were treated as continuous and standardized to have mean 0 and variance 1. In our simulations, BMI values were generated as follows:

$$BMI_M = -0.116\, Smoking_M - 0.129\, SEP_M + 0.4\, U_2 + \varepsilon_M$$

$$BMI_P = -0.105\, Smoking_P - 0.088\, SEP_P + 0.4\, U_2 + \varepsilon_P$$

As with smoking, the confounding factor $U_2$ represents the correlation between paternal and maternal BMI, and its effect of 0.4 was set to ensure that paternal and maternal BMI were correlated with the same correlation as in ALSPAC. The effects of smoking and SEP on BMI were obtained from linear regression analyses in ALSPAC. Finally, the error terms were distributed as $\varepsilon_M \sim N(0.201, 0.911^2)$ and $\varepsilon_P \sim N(0.165, 0.918^2)$, to ensure that BMI values were normally distributed with mean 0 and variance 1.

Birthweight data were simulated according to

$$Birthweight = 0.055\, BMI_P + 0.141\, BMI_M - 0.155\, Smoking_P - 0.309\, Smoking_M + \varepsilon_{BW} \quad (1)$$

where again, the coefficients of parental BMI and smoking were set equal to the values observed in ALSPAC and the error term $\varepsilon_{BW}$ was distributed as $\varepsilon_{BW} \sim N(0.168, 0.972^2)$, to ensure that birthweight values were normally distributed with mean 0 and variance 1.

Paternal participation was treated as a binary variable, and its values were generated from a logistic regression model with probability of participation:

$$logit\ P(Participation = 1) = \alpha + \beta_1 SEP_P + \beta_2 SEP_M + \beta_3 BMI_P + \beta_4 Smoking_P$$

Here, $\beta_2$ denotes the effect of maternal SEP on paternal participation; this parameter could be estimated in the ALSPAC dataset and was set equal to $\beta_2 = 0.324$. On the other hand, the selection effects $\beta_1, \beta_3, \beta_4$ could not be estimated due to the unavailability of smoking, BMI and SEP data for non-participating partners. In our simulations we varied the values of these parameters and considered eleven different scenarios, which are summarized in Supplementary Table 1. Finally, the intercept $\alpha$ was tuned to ensure that paternal participation remained approximately at ALSPAC levels (76.7%), regardless of the values of the other parameters.

We now describe the eleven simulation scenarios we considered. Scenario 1 was created by computing the effects of maternal BMI and smoking on paternal participation in ALSPAC and setting the paternal effects to be equal to the observed maternal effects, while the paternal SEP effect was estimated using paternal SEP data provided by the mothers about their partners. The values of the associations between these parental traits and birthweight in ALSPAC were estimated from separate models. In Scenarios 2-6, we assumed no direct effect of smoking on participation and varied the values of the SEP-Participation and BMI-Participation effects. We used logarithmic values to allow the coefficients to be easily interpreted as odds ratios. We considered odds ratios of 1.5, 2, 5, 10 and 20 for the increase in the odds of partner participation per unit increase in the values of SEP and (standardized) BMI. These values were selected for exploratory purposes, as they represent different degrees of association between each paternal trait and participation. In Scenarios 7-11 we also allowed for a Smoking-Participation effect and varied its value along these of the other two selection effects (but in the opposite direction, since smoking decreases the likelihood of participation).

We did not implement a factorial design, as the number of simulation scenarios would be too large to handle (a factorial design with the parameter values of Supplementary Table 1 would have resulted in $6 \times 6 \times 7 = 252$ different scenarios). Instead, we simply assumed that the three selection effects $\beta_1, \beta_3, \beta_4$ were of similar strength (except in scenario 1).

The simulation experiment was repeated 1000 times for each scenario.

*Estimand*

The quantities of interest in our simulation study were the effects of maternal and paternal smoking on offspring birthweight, adjusted for SEP. Since a perfect indicator of overall health would be practically impossible to obtain in ALSPAC, our analyses were not adjusted for parental overall health but only for SEP. This meant that our estimands represent the total effects of smoking on offspring birthweight for each parent (both direct and mediated by parental general health). The true values for these total effects in our simulation study were $-0.3254$ for maternal and $-0.1608$ for paternal smoking respectvely.

*Methods*

To estimate the effects of interest, we fitted linear regression models with offspring birthweight as the outcome and maternal and paternal smoking as covariates. We also adjusted for parental SEP, as is likely to be the case in real-data analyses (note that parental SEP acts as a confounder between smoking and birthweight when not adjusting for general health). This means that the model from which we estimated the effects of smoking on birthweight was

$$Birthweight \sim Smoking_M + Smoking_P + SEP_M + SEP_P$$

We did not implement any methods to correct for selection bias (such as inverse probability weighting). Our objective in this simulation study was simply to assess the magnitude of bias.

*Performance measures*

For each of the 1000 replications, we computed the regression coefficients for the association of maternal and paternal smoking with offspring birthweight, as well as their standard errors and 95% confidence intervals. We then averaged these across the 1000 replications and obtained five performance measures with which to assess the impact of selection bias in our simulation study: mean bias in parameter estimates for the smoking-birthweight associations, mean relative bias, average standard errors, the empirical coverage of 95% confidence intervals, and bias-eliminated coverage. The relative bias of an estimator $\hat{\theta}$ for a parameter $\theta$ is defined as $RB(\theta, \hat{\theta}) = \frac{\hat{\theta}-\theta}{\theta}$. Empirical coverage was computed as the proportion of replications of our simulation study in which the estimated 95% confidence interval contained the true parameter value. Bias-eliminated coverage was defined analogously as the proportion of replications of the simulation in which the estimated 95% confidence interval contained the average estimate. This performance measure can be used to assess whether any reduction in the coverage of confidence intervals is due to bias or poor calibration of standard errors, and hence whether selection bias only affects the parameter estimates themselves or whether it also biases the estimated standard errors.

*Simulation results*

The results of our simulation experiment were presented in Figure 5 of the main part of the paper and are also reported in Supplementary Table 2. As discussed in the paper, there was little evidence of selection bias in maternal effect estimates, under any of our simulation scenarios. For paternal effects, we observed selection bias mainly in Scenarios 9-11, in which all three paternal variables (SEP, smoking and general health) were strongly associated with participation. However, the magnitude of bias was fairly small, with a worst-case relative bias of -12.6% for paternal effect estimates (in Scenario 11 – note that a negative sign indicates upward bias, since the parameter to be estimated was negative).The bias also affected the coverage of 95% confidence intervals, with reduced coverage being observed in Scenarios 9-11, although the small magnitude of bias meant that confidence intervals for the paternal effects still contained the true value more often than not, as illustrated in Figure 5. On the other hand, selection bias did not seem to affect the model-based standard errors, with similar standard error estimates obtained in all simulation scenarios. Accordingly, bias-eliminated coverage remained at nominal levels.

*Assessing bias with Monte Carlo errors*

To better assess which simulation scenarios provided evidence of selection bias, we computed Monte Carlo errors for the empirical coverage of 95% confidence intervals. If selection bias was present in our simulations, we would expect parameter estimates to deviate from their true values and the coverage of 95% confidence intervals to fall below the nominal level of 95%. In the absence of selection bias, some variability in the results should still be expected due to chance. Monte Carlo errors can be used to quantify how much variability should be expected by chance alone. Since we performed 1000 iterations, the Monte Carlo error for coverage in our simulations under the assumption of no selection bias was $\sqrt{0.95 * 0.05/1000} \approx 0.69\%$, which would suggest a 95% confidence interval of (93.65%, 96.35%) for the coverage figures. However, between our two different estimands (paternal and maternal effects) and eleven scenarios, we are performing 22 different simulations. We therefore adjusted for multiple testing and used the $(1-\frac{0.05/22}{2})$-th quantile of the $N(0,1)$ distribution, instead of the $(1-\frac{0.05}{2})$-th quantile, which yielded a confidence interval of approximately (92.9%, 97.1%). In scenarios where the empirical coverage was outside this range, we concluded that there is evidence of selection bias. This happened in the three simulation scenarios mentioned earlier (9-11), and the effect of paternal smoking on offspring birthweight was overestimated in these scenarios.

*Additional Simulations*

Our previous simulation indicated that selection bias occurred when paternal general health was not accounted for. In that simulation, paternal general health acted as a common cause (confounder) of paternal participation and birthweight. The presence of such a common cause between participation and the outcome can result in selection bias: in the DAG of Figure 2, conditioning on paternal participation opens the path "Paternal smoking → Paternal participation → Paternal Health → Birthweight". According to the theory of d-separation, adjusting for paternal overall health would be enough to block this pathway and eliminate the bias. On the other hand, the presence of additional common causes between partner participation and birthweight, apart from paternal health, could increase the impact of selection bias. Such common causes would also need to be adjusted for in order to eliminate the bias completely, but this can be difficult in real-data applications where one cannot be sure that all the relevant common causes have been identified.

To further explore the effects of selection-outcome confounding on selection bias, we generated two additional sets of simulations. In the first set, we used the same data-generating model as described above but assumed that an accurate measure of parental general health (BMI) is available and our analysis for the effects of smoking on birthweight can also be adjusted for parental general health. The purpose of this simulation was to illustrate that when all common causes of participation and the outcome are accounted for, selection bias will not occur. Since our analysis was now adjusted for parental health, the estimands of interest were the direct effects of maternal and paternal smoking on birthweight, not mediated by parental health. To estimate the direct effects, we fitted the model

$$Birthweight \sim Smoking_M + Smoking_P + BMI_M + BMI_P + SEP_M + SEP_P$$

using BMI as a proxy for general health.

Results of this additional simulation are reported in Supplementary Table 3. Selection bias was not detected in any of our eleven simulation scenarios, for either maternal or paternal effect estimates. This is in line with the relevant theory and suggests that selection bias will not arise when all common causes of partner participation and the outcome (birthweight) have been adjusted for.

In our second set of further simulations, we included an additional unobserved confounder (common cause) of paternal participation and birthweight. The data generating model of our ADEMP design was modified to include an outcome-selection confounder $U_{BW}$. Specifically, we modified our model for birthweight to be

$$Birthweight = 0.055\,BMI_P + 0.141\,BMI_M - 0.155\,Smk_P - 0.309\,Smk_M + U_{BW} + \varepsilon'_{BW}$$

The amount of variation in birthweight explained by the variable $U_{BW}$ was specified so that the correlation between birthweight and paternal participation in the simulations was

$Cor(Birthweight, U_{BW}) = 0.0476$, which is the value observed in ALSPAC. The error term $\varepsilon'_{BW}$ was again specified so that birthweight measurements correspond to Z scores (i.e. have zero mean and unit variance). Likewise, the model for an individual's probability of participation was modified to be

$$logit\ P(Participation = 1) = \alpha + \beta_1 SEP_P + 0.324\,SEP_M + \beta_3 BMI_P + \beta_4 Smoking_P + U_{BW}$$

with the selection effects $\beta_1, \beta_3, \beta_4$ varied according to the eleven simulation scenarios described in Supplementary Table 1 and the intercept $\alpha$ tuned to ensure that the proportion of participating fathers was 76.7%. We estimated the effects of paternal and maternal smoking on birthweight adjusted for SEP but not general health, meaning that the estimand of interest was the total (not the direct) effect of smoking on birthweight, as in our initial simulation.

The results of this additional simulation experiment are reported in Supplementary Table 4. Selection bias was observed in several simulation scenarios for paternal effects. Maternal effect estimates were still unbiased, since there was no association between maternal smoking and paternal participation once paternal smoking had been taken into account. The magnitude of bias will likely depend on the strength of confounding, but in any case it was stronger than in our original simulations in Supplementary Table 2, since there are now two common causes of paternal participation and offspring birthweight that were not adjusted for. In addition, the magnitude of bias depends on the selection effects; hence, it was again stronger in simulations when paternal smoking directly affected participation and the values assigned to the selection effects were large. We also note that bias was detected even in Scenario 1, where associations between maternal traits and paternal participation in ALSPAC were used as a substitute for paternal selection effects.

Nevertheless, it should be noted that the magnitude of bias remained small in all scenarios. In the worst-case scenario among our simulations (scenario 11 for paternal effects) the average absolute bias was 0.029 while the corresponding relative bias was -18.1%. In all simulation scenarios, the biased estimates were less than two standard deviations away from the true value of the parameter they aimed to estimate. And finally, our analysis was able to (correctly) reject the null hypothesis of no effect of (paternal or maternal) smoking on birthweight in all scenarios, despite the presence of bias.

*Limitations of our simulation study*

Limitations of this simulation study include the fact that it was parametrized using observational associations, which may or may not be the same as causal effects, as well as the fact that the use of BMI as a proxy for general health may not always be accurate due to the fact that its effects are often non-linear. For example, while we would expect the effect of SEP on general health to be positive, the observed SEP-BMI associations in ALSPAC were negative since high-SEP individuals typically have moderate but not very high BMI. In addition, our simulations relied on the causal diagram of Figure 2 being realistic. Finally, we did not consider scenarios containing interactions between the variables in our analysis, in their effects on paternal participation. Such interactions can often exacerbate the effects of selection bias.

Nevertheless, subject to these limitations, we conclude that although selection bias may arise in an ALSPAC-based analysis of the association between paternal and maternal smoking habits and offspring birthweight, it is unlikely to have a large impact on the results of such an analysis.

**Supplementary Materials**

Supplementary Table 1. Simulation scenarios. Scenarios varied in terms of their parameterized effect of partner SEP, general health (proxied by BMI), and smoking on partner participation. The reported values represent odds ratios of increase in paternal participation per unit increase in each variable (general health/BMI was standardized to variance 1, so “unit increase” in general health actually refers to one standard deviation increase). As odds ratios, they correspond to the values of $\exp(\beta_1)$, $\exp(\beta_3)$ and $\exp(\beta_4)$ in our simulation design respectively ($\beta_2$ denotes the effect of maternal SEP on paternal participation, which could be estimated in ALSPAC and was set equal to 0.324).

| Scenario | Parameter Value | | |
|---|---|---|---|
| | SEP | General Health | Smoking |
| 1 | $\exp(0.46)$ | 1 | $\exp(-0.81)$ |
| 2 | 1.5 | 1.5 | 1 |
| 3 | 2 | 2 | 1 |
| 4 | 5 | 5 | 1 |
| 5 | 10 | 10 | 1 |
| 6 | 20 | 20 | 1 |
| 7 | 1.5 | 1.5 | 2/3 |
| 8 | 2 | 2 | 1/2 |
| 9 | 5 | 5 | 1/5 |
| 10 | 10 | 10 | 1/10 |
| 11 | 20 | 20 | 1/20 |

Supplementary Table 2: Absolute bias, relative bias, model-based standard errors, empirical coverage of 95% confidence intervals and bias-eliminated coverage for the effects of paternal and maternal smoking on offspring birthweight in our simulation study. Estimates obtained from models adjusted for parental SEP but not general health.

| Scenario | Paternal/Partner Effects | | | | | Maternal Effects | | | | |
|---|---|---|---|---|---|---|---|---|---|---|
| | Abs Bias | Rel Bias | Std Error | 95% Cov | B-E Cov | Abs Bias | Rel Bias | Std Error | 95% Cov | B-E Cov |
| 1 | 0.001 | -0.5% | 0.021 | 95.5% | 95.3% | 0.000 | -0.1% | 0.023 | 95.9% | 95.7% |
| 2 | 0.001 | -0.7% | 0.021 | 94.0% | 94.5% | -0.001 | 0.2% | 0.023 | 94.7% | 95.0% |
| 3 | -0.001 | 0.6% | 0.021 | 94.9% | 94.8% | 0.001 | -0.3% | 0.023 | 95.6% | 95.2% |
| 4 | 0.000 | -0.1% | 0.021 | 95.5% | 95.4% | 0.001 | -0.2% | 0.023 | 93.9% | 93.9% |
| 5 | 0.002 | -1.3% | 0.021 | 95.1% | 94.9% | 0.000 | 0.0% | 0.023 | 94.8% | 94.8% |
| 6 | 0.001 | -0.6% | 0.021 | 93.8% | 94.0% | 0.000 | -0.1% | 0.023 | 94.9% | 95.0% |
| 7 | 0.002 | -1.4% | 0.021 | 96.3% | 96.3% | 0.000 | 0.1% | 0.023 | 96.3% | 96.5% |
| 8 | 0.006 | -3.4% | 0.021 | 93.9% | 95.2% | 0.001 | -0.2% | 0.023 | 93.5% | 93.5% |
| 9 | 0.013 | -8.1% | 0.021 | 90.6% | 95.1% | 0.002 | -0.8% | 0.023 | 94.4% | 94.1% |
| 10 | 0.019 | -11.5% | 0.021 | 85.1% | 94.4% | 0.000 | 0.1% | 0.024 | 95.1% | 94.9% |
| 11 | 0.020 | -12.6% | 0.021 | 83.6% | 94.4% | 0.000 | -0.1% | 0.024 | 95.1% | 95.3% |

Supplementary Table 3: Absolute bias, relative bias, model-based standard errors, empirical coverage of 95% confidence intervals and bias-eliminated coverage for the effects of paternal and maternal smoking on offspring birthweight in our simulation study. Estimates obtained from models adjusted for both parental SEP and general health.

| Scenario | Paternal/Partner Effects | | | | | Maternal Effects | | | | |
|---|---|---|---|---|---|---|---|---|---|---|
| | Abs Bias | Rel Bias | Std Error | 95% Cov | B-E Cov | Abs Bias | Rel Bias | Std Error | 95% Cov | B-E Cov |
| 1 | 0.001 | -0.5% | 0.021 | 94.7% | 95.0% | 0.000 | -0.1% | 0.023 | 95.7% | 95.7% |
| 2 | 0.001 | -0.7% | 0.021 | 94.6% | 94.9% | -0.001 | 0.2% | 0.023 | 94.6% | 94.7% |
| 3 | -0.001 | 0.9% | 0.021 | 94.5% | 94.5% | 0.001 | -0.3% | 0.023 | 95.0% | 94.9% |
| 4 | -0.001 | 0.9% | 0.021 | 94.7% | 94.9% | 0.001 | -0.2% | 0.023 | 93.7% | 94.2% |
| 5 | 0.001 | -0.4% | 0.021 | 95.5% | 95.7% | 0.000 | 0.0% | 0.023 | 95.0% | 95.1% |
| 6 | -0.001 | 0.7% | 0.021 | 94.6% | 94.5% | 0.000 | -0.1% | 0.023 | 95.0% | 94.9% |
| 7 | 0.000 | -0.1% | 0.021 | 95.6% | 95.5% | 0.000 | 0.1% | 0.023 | 96.0% | 96.2% |
| 8 | 0.000 | -0.3% | 0.021 | 94.9% | 94.9% | 0.000 | -0.1% | 0.023 | 93.5% | 93.4% |
| 9 | -0.001 | 0.7% | 0.021 | 95.0% | 95.4% | 0.002 | -0.7% | 0.023 | 94.2% | 94.2% |
| 10 | 0.000 | -0.2% | 0.021 | 94.8% | 94.9% | -0.001 | 0.2% | 0.023 | 95.2% | 95.1% |
| 11 | -0.001 | 0.3% | 0.021 | 93.9% | 94.0% | 0.000 | -0.1% | 0.023 | 94.8% | 94.9% |

Supplementary Table 4: Absolute bias, relative bias, model-based standard errors, empirical coverage of 95% confidence intervals and bias-eliminated coverage for the effects of paternal and maternal smoking on offspring birthweight in simulations with an additional common cause of paternal/partner participation and birthweight. Estimates obtained from models adjusted for parental SEP but not general health.

| Scenario | Paternal/Partner Effects | | | | | Maternal Effects | | | | |
|---|---|---|---|---|---|---|---|---|---|---|
| | Abs Bias | Rel Bias | Std Error | 95% Cov | B-E Cov | Abs Bias | Rel Bias | Std Error | 95% Cov | B-E Cov |
| 1 | 0.012 | -7.6% | 0.021 | 91.4% | 95.6% | -0.001 | 0.2% | 0.023 | 95.4% | 95.3% |
| 2 | 0.000 | -0.2% | 0.021 | 94.4% | 94.2% | 0.001 | -0.2% | 0.023 | 93.5% | 93.4% |
| 3 | 0.002 | -1.1% | 0.021 | 94.5% | 94.9% | -0.001 | 0.3% | 0.023 | 94.6% | 95.0% |
| 4 | 0.003 | -1.9% | 0.021 | 95.0% | 95.2% | 0.000 | -0.1% | 0.023 | 95.1% | 95.1% |
| 5 | 0.004 | -2.3% | 0.021 | 94.9% | 95.3% | 0.000 | -0.1% | 0.023 | 94.7% | 94.8% |
| 6 | 0.003 | -1.7% | 0.021 | 95.4% | 95.7% | -0.001 | 0.2% | 0.023 | 94.8% | 94.6% |
| 7 | 0.009 | -5.7% | 0.021 | 93.5% | 94.9% | 0.001 | -0.3% | 0.023 | 94.5% | 94.5% |
| 8 | 0.015 | -9.4% | 0.021 | 88.8% | 95.0% | -0.001 | 0.5% | 0.023 | 95.1% | 95.1% |
| 9 | 0.026 | -16.2% | 0.021 | 77.5% | 95.4% | 0.000 | 0.0% | 0.023 | 95.2% | 95.1% |
| 10 | 0.028 | -17.3% | 0.021 | 74.0% | 94.5% | 0.001 | -0.3% | 0.024 | 93.9% | 94.2% |
| 11 | 0.029 | -18.1% | 0.021 | 71.4% | 95.6% | 0.000 | 0.1% | 0.024 | 94.8% | 94.8% |
